# Initial Clinical Outcomes from NOCD Digital Behavioral Health Treatment of Obsessive-Compulsive Disorder Using Exposure and Response Prevention

**DOI:** 10.1101/2021.01.18.20173633

**Authors:** Jamie D. Feusner, Patrick B. McGrath, Ted Faneuff, Stephanie Lonsway, Reza Mohideen, Larry Trusky, Stephen M. Smith

**Affiliations:** NOCD, LLC, Chicago, IL, 60611, USA; Department of Psychiatry and Biobehavioral Sciences, University of California Los Angeles, Los Angeles, CA 90095, USA

## Abstract

Effective first-line treatments for obsessive-compulsive disorder (OCD) include exposure and response prevention (ERP) therapy. Despite extensive evidence of its efficacy in clinical studies and real-world samples, ERP is still underutilized as a treatment, likely due to access to care barriers such as the availability of adequately trained ERP therapists, geographical location, time, and cost. NOCD has created a digital behavioral health treatment for OCD using ERP delivered via teletherapy and between-session support. We examined preliminary treatment outcomes in a large naturalistic sample of 2069 adults, children, and adolescents with a primary OCD diagnosis. Treatment consisted of twice-weekly live teletherapy ERP for three weeks, followed by six weeks of once-weekly brief teletherapy check-ins. Assessments were conducted at baseline, after completion of three weeks of twice-weekly sessions, and at the end of the six weeks of brief check-ins. Treatment resulted in significant improvements, with a 45% mean reduction in OCD symptoms and a 71% response rate (≥35% reduction in OCD symptoms).Treatment also resulted in a significant, 43% reduction in depression, a 49% reduction in anxiety, and a 35% reduction in stress symptoms. Quality of life improved by a mean of 35%. The mean duration of treatment was approximately 11 weeks, and the total therapist time was approximately 11 hours, which is less than half the total time compared with standard once-weekly outpatient treatment. In sum, in this preliminary sample, NOCD’s treatment model for OCD, delivered in a readily-accessible format for patients, has demonstrated to be effective and efficient.

## Background

Obsessive-compulsive disorder (OCD) is a prevalent and often disabling psychiatric disorder, affecting 2.3% of individuals during their lifetimes (Ruscio et al. 2008). Exposure and response prevention (ERP), a type of cognitive-behavioral therapy, is effective for OCD and is considered a first-line treatment ([American Psychiatric Association Practice Guidelines](https://psychiatryonline.org/pb/assets/raw/sitewide/practice_guidelines/guidelines/ocd.pdf), [UK NICE Clinical Guideline](https://www.nice.org.uk/guidance/cg31)). However, ERP requires specialty-trained therapists and thus is not readily available to everyone with OCD because of cost and geographical limitations (O’Neill and Feusner 2015). Moreover, ERP, when delivered in its most common format - once weekly outpatient therapy - typically requires over 25 hours of therapist time per patient (Foa, Yadin, and Lichner 2012) to achieve meaningful results, which could take six or more months.

To address the challenges of this evidence-based treatment of 1) barriers to access, and 2) associated cost and time, NOCD has developed a digital behavioral health treatment program for ERP using video teletherapy. Remote ERP for OCD, delivered by video or telephone, has been demonstrated in a meta-analysis to significantly improve OCD symptoms with large effect sizes (Hedges g from 1.21-2.77) (Wootton 2016). Head-to-head comparisons with in-person treatment in adults and adolescents show no meaningful differences in outcome (Lovell et al. 2006; Turner et al. 2014). One of several vital advantages of remote treatment is that therapists can readily interact with patients in the settings that might trigger their symptoms the most, such as in the home, to administer in-session exposures that otherwise could be difficult or impossible to reproduce in the office. Moreover, as of 2018, 81% of adults in the United States owned smartphones (Pew Research Center, 2019), which grows yearly.

NOCD’s treatment model is based on a clinical trial conducted by researchers at Columbia University Medical Center and the New York State Psychiatric Institute (Gershkovich et al. 2020) that used the NOCD app integrated with brief in-person therapy and remote check-ins. This study tested a treatment protocol designed to minimize therapist time (mean 402±91 minutes - under 7 hours - total per patient) while increasing therapy intensity compared with once-weekly ERP sessions. In this study, only 17.8% of potentially eligible participants declined to enroll, and there were high satisfaction ratings: 68.2% were “very” and 31.8% “mostly” satisfied with services received. The treatment resulted in a mean reduction in OCD symptoms of 38.9%, with a response rate of 52% (≥35% reduction in OCD symptoms). We designed a similar treatment for NOCD to treat OCD patients in the general community based on this overall structure, with two primary modifications: a) to further improve accessibility all sessions were conducted remotely with video teletherapy, and b) to provide additional support, enhance adherence, and potentially improve efficacy every patient had access to between-session contact with his/her/their therapist via messaging and support from an online OCD community.

## Methods

### Treatment Model

The NOCD treatment model consists of an initial diagnostic assessment followed by twice-weekly 60-90 minute ERP video sessions for 2-3 weeks. After this, patients then had six weeks of once-weekly 30-minute video “check-in” sessions to guide ongoing ERP “homework” assignments conducted by the patients. Also, between sessions, all patients had access to as-needed asynchronous text messaging with their therapists to obtain guidance with exposures and response prevention. Patients also had 24 hours per day, seven days per week access to the online NOCD community, consisting of a (monitored) forum of individuals around the world self-identified as having OCD, providing support and advice through online postings.

### Assessments

To measure OCD symptoms, we used the patient-rated Dimensional Obsessive-Compulsive Scale (DOCS) (Abramowitz et al. 2010) and the clinician-rated Diagnostic Interview for Anxiety, Mood, and OCD and Related Neuropsychiatric Disorders (DIAMOND) OCD severity scale (Tolin et al. 2018). To measure depression, anxiety, and stress symptoms, we used the DASS-21 scale (Henry and Crawford 2005). To measure quality of life, we used the Quality of Life Enjoyment and Satisfaction Questionnaire -- Short Form (QLES-Q) (Endicott et al. 1993). Assessments were conducted at baseline at the time of the initial diagnostic assessment, at treatment midpoint (4 weeks for most patients), and the endpoint (8-12 weeks for most patients).

### Statistical Analysis

Data analysis was conducted using linear mixed models with assessment number as a fixed factor, patient as a random factor, and DOCS and DIAMOND OCD severity scores as the dependent variables. Secondary outcome analyses for the QLES-Q and the DASS-21 subscales of depression, anxiety, and stress were analyzed using the same model. We analyzed data using SPSS version 26.0.0.0.

## Results

### Sample

In this ongoing treatment sample, as of November 8, 2020, n=2069 adults, adolescents, and children have received at least an initial evaluation and received a primary diagnosis of OCD. (n=155 of the total sample were children or adolescents). At the time of analysis, n=1163 had completed at least the midpoint assessment, and n=620 had completed the initial, midpoint, and endpoint assessments. The mean treatment duration was 11.02±2.90 weeks, the mean number of therapist sessions was 13.66±2.68, and the mean number of therapist hours was 10.79±1.79.

### OCD symptom results

NOCD treatment, by endpoint, resulted in a significant decrease in patient-rated OCD symptoms (DOCS scores) (F_2,1742.39_=719.19, p<.001; Hedges g = 1.16: “large” effect size). It also resulted in a significant decrease in clinician-rated OCD symptoms (DIAMOND OCD scores) (F_2,2164.17_=1059.54, p<.001; Hedges g = 1.64: “large” effect size). By midpoint, there were also statistically significant improvements, with an average of 27.78% reduction in patient-rated OCD symptom severity (t_977_=26.49, p<.001) and a 22.03% reduction in clinician-rated OCD severity (t_1115_=30.82, p<.001).

By endpoint, treatment has resulted in an average of 45.25% decrease in patient-rated OCD severity (DOCS) and an average of 38.26% decrease in clinician-rated OCD severity (DIAMOND). Further, 71.0% of those who have completed treatment thus far meet criteria as “responders,” traditionally defined as a ≥35% reduction in OCD symptoms, based on DOCS scores, and 53.6% based on DIAMOND OCD scores.

**Fig. 1.**
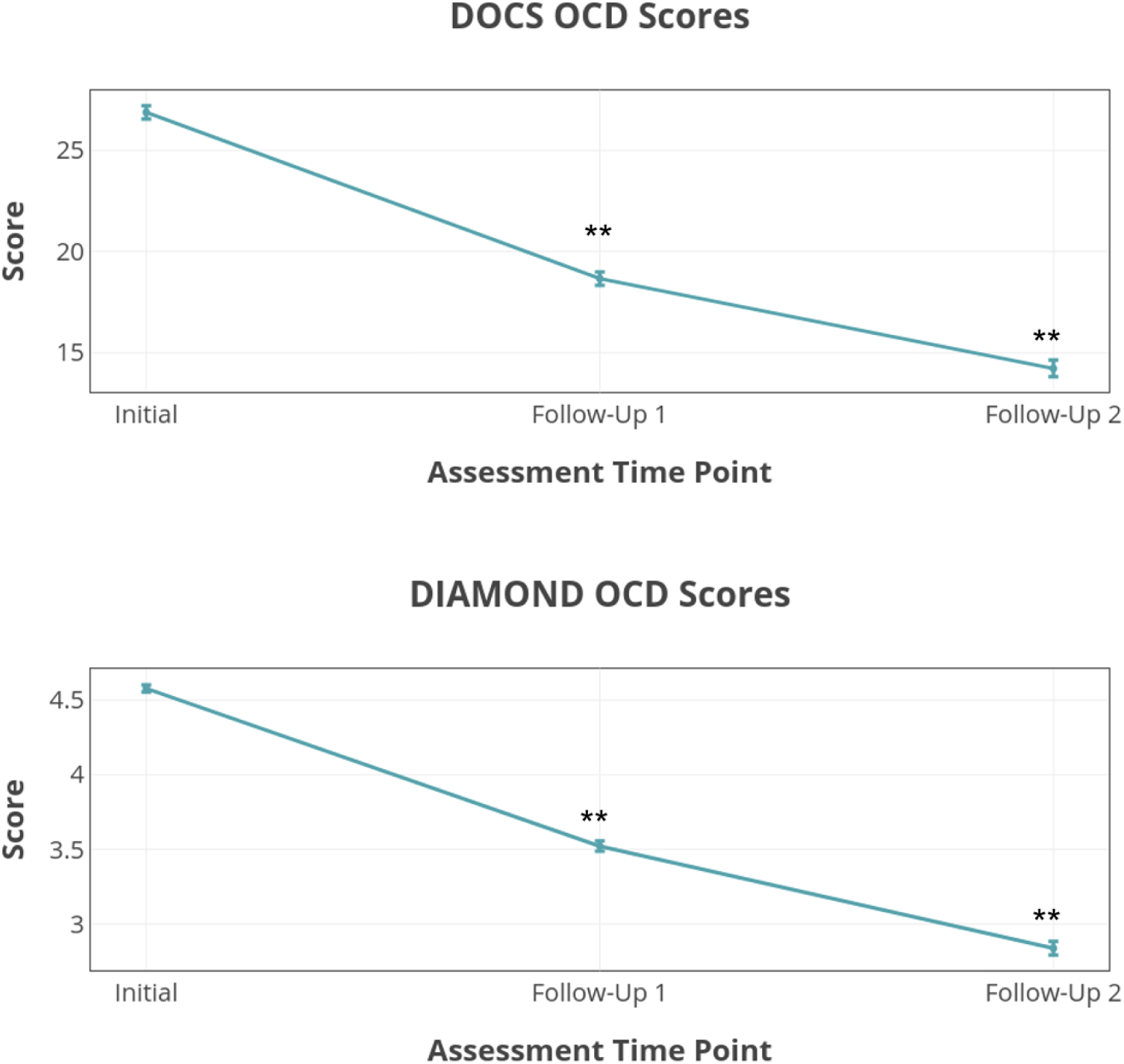
Improvement in patient-rated OCD symptoms as assessed by the Dimensional Obsessive-Compulsive Scale (DOCS), top; and by clinician-rated DIAMOND OCD Scale, bottom. Error bars indicate standard error of the mean. ∗∗p<.001 compared with initial scores.

### Depression, Anxiety, Stress, and Quality of Life Results

- By endpoint:
- Depression scores decreased by an average of 43.27% (g=.93, large effect size).
- Anxiety scores decreased by an average of 49.19% (g=1.14, large effect size).
- Stress scores decreased by an average of 35.11% (g=1.08, large effect size).
- Quality of life (QLES-Q) increased by an average of 34.65% (g=1.22, large effect size).
- All improvements were statistically significant (p<.001).

## Discussion

From this preliminary analysis of patients with OCD, digital teletherapy treatment with NOCD using ERP has resulted in significant improvement in OCD symptoms. Mean patient-rated symptom reduction was approximately 45%, with a 70% response rate. In addition to these promising results thus far in reducing OCD symptoms, NOCD treatment also resulted in improvements in other comorbid symptoms common in OCD: depression, anxiety, and stress. Therefore, a single focused OCD treatment can result in an overall reduction of multiple disabling and distressing symptoms. It also resulted in a significant 35% improvement in quality of life in just 11 weeks. This is substantial in the context of the fact that OCD is a chronic illness that individuals suffer on average for 11 years before getting treatment (Pinto et al. 2006). Long term follow-up data at 3, 6, 9, and 12 months post-treatment are currently being collected.

This treatment model for NOCD was developed to provide an evidence-based form of treatment, ERP, in a manner that is readily accessible for patients (teletherapy) and is efficient in terms of total therapist time. Additional support for patients between sessions is provided through patient-therapist messaging and 24-hour access to NOCD’s online support community. The clinician-rated OCD symptom results obtained to date are similar to the Columbia University study results (Gershkovich et al. 2020), despite the current results being from a “real-world” clinical sample rather than a research sample with more selective inclusion and exclusion criteria. Direct comparisons are limited by the fact that NOCD used the DIAMOND OCD severity scale, whereas the Columbia study used the Yale-Brown Obsessive-Compulsive Scale (Goodman et al. 1989). Also, NOCD used the patient-rated DOCS scale, yet a patient-rated scale was not used in the Columbia study.

These current results demonstrate not only the magnitude of the effect of this treatment protocol on OCD and comorbid symptoms but also its efficiency in terms of cost and time savings. The current results were achieved in less than half the total therapist time than traditional outpatient ERP (Foa, Yadin, and Lichner 2012) and with a treatment course that was 56% shorter than the duration of typical outpatient treatment. This has the potential for substantial cost savings for patients and third-party payors (e.g. health insurers).

## Conclusions

In sum, NOCD treatment in this preliminary sample shows promising results for effective and efficient treatment for OCD on a large scale and is delivered in a readily-accessible format for patients. Further, NOCD treatment represents a substantial cost savings for patients and third-party payors over both traditional weekly outpatient ERP.

## Data Availability

Data may be available upon request.

